# Professionals’ views on the mental health problems and vulnerability of children and young people during the early phase of the COVID-19 pandemic

**DOI:** 10.1101/2021.04.26.21256103

**Authors:** Julian Edbrooke-Childs, Angelika Labno, Melissa A. Cortina, Anna Gilleard, Daniel Hayes, Yeosun Yoon, Christian Dalton-Locke, Sonia Johnson, Alan Simpson, Norha Vera San Juan, Ellie Brooks-Hall, Mental Health Policy Research Unit

## Abstract

The COVID-19 pandemic caused major disruptions to everyday life for children and young people. The aim of this study was to examine professionals’ views on the mental health problems and vulnerabilities of children and young people during the early phase of the COVID-19 pandemic. We conducted a secondary analysis of an online survey completed by mental health professionals in the UK between 22 April 2020 and 12 May 2020. The final sample was *N* = 601 professionals who at least partly worked with children and young people. Quantitative and qualitative survey data showed that professionals were concerned about young people experiencing economic disadvantage and young people from minority ethnic groups, as pre-existing social inequalities resulted in increased risk of infection and reduced access to physical and mental health care. Professionals were concerned about young people with little family support and young people at risk of relapse or deterioration in mental health, reporting the exacerbation of pre-existing mental health difficulties and challenging behaviours. Further research, involving young people as researchers, is needed to explore the continued impact for children and young people, and their families, including in comparison to their experiences before the pandemic.

## 1. Introduction

The COVID-19 pandemic caused major disruptions to everyday life for children and young people. School closures and periods of confinement decreased the amount of social interaction, removed access to support services, and increased concerns about the future. Evidence suggests the pandemic triggered a deterioration in mental health in children and young people globally (Cortina et al., 2020; Cortina et al., 2020; Gilleard et al., 2020). A study in the United Kingdom (UK) found significantly increased rates of mental distress in young people aged 16-24 years during the pandemic period compared with rates projected for the same period (Pierce et al., 2020). Another study in the United States (US) similarly found a higher prevalence of clinical depression, anxiety, and post-traumatic stress syndrome in young adults during the early phase of the pandemic (Liu et al., 2020).

Emerging evidence shows how the social, economic, and health impacts of the pandemic are exacerbating existing socio-economic inequalities for children and young people across the world (Institute of Healthy Equity, 2020). More economically vulnerable families may not have the financial supports to protect children’s health and wellbeing. School closures have amplified the divide between the wealthy and the poor, and the impact can be seen on learning and food security. Children from higher-income families reported having more access to resources for home learning and spent more time on home learning (Andrew et al., 2020). These gaps in education are expected to widen educational inequalities over time.

The impact of pandemic-related stress on families has been viewed through the lens of ‘psychosocial stress contagion’ (Liu & Doan, 2020), which posits stress in one domain of life can spill over into other areas, affecting both parents and children. For example, new stressors on job demands or personal finances can compromise parenting abilities to provide adequate childcare. Children, who are absorbing these spill-over effects from caregivers in addition to navigating their own stressors, may have more trouble regulating their emotions. A study showed that as the number of hardships increased (e.g. job loss, income reduction), the psychological wellbeing of parents and children worsened (Gassman-Pines et al., 2020).

Young people who are known to social care have experienced reduced wraparound supports, placements, or contact with birth parents and relatives during the pandemic. Kinship care and foster care arrangements have experienced increased strain as they are providing more care for children during school closures with less opportunities for respite (Galvin & Kaltner, 2020). Similarly, young people with caring responsibilities at home have reported additional stressors under lockdown and less time for themselves, as well as reduced access to support from wider family, peers, teachers, and health and social care services (Benson-Allott, 2020; Blake-Holmes, 2020). However, some young carers have reported feeling relief from previous pressures during lockdown, such as social anxiety and bullying connected with school time (Children’s Commissioner for Wales, 2020). Young people in residential care or temporary accommodation are thought to be at particular risk of significant emotional stress and health issues as they may be forced to relocate or return to households without adequate supports as a result of COVID-19 (Goldman et al., 2020).

Research from the early period of the COVID-19 pandemic indicates children and young people with pre-existing physical and mental health conditions may be particularly impacted by the pandemic. Mental health concerns appeared to be exacerbated in a sample of young people with pre-existing physical health concerns, particularly those with allergies or asthma (Hawke et al., 2020). Young people with a pre-existing mental illness have also reported significant levels of depression, anxiety and PTSD in the US (Liu et al., 2020). Another group identified as particularly vulnerable is young people with neurodevelopmental disorders such as autism spectrum disorder (ASD), attention-deficit/hyperactivity disorder (ADHD), and obsessive-compulsive disorder (OCD). A UK study showed a high prevalence of emotional and behavioural difficulties in a group of children and young people with ASD or ADHD, and especially in those with a comorbid diagnosis (Nonweiler et al., 2020). Young people with OCD and eating disorders have reported relapse or an increase in symptom severity during the pandemic (Graell et al., 2020; Tanir et al., 2020). Risk factors for relapse or the worsening of symptoms include the lack of structure and routines resulting from necessary school closures, reduced access to services, and physical health and contamination concerns.

Given the generalised recognition of rising need and the widespread restructuring of children and young people’s mental health services, mental health professionals working on the frontline are well positioned to elucidate some of these mental health challenges they are observing in clinical practice. Richly descriptive research is needed to better understand the emerging mental health needs of children and young people during the pandemic (Das, 2020).

The aim of the present article is to examine professionals’ views on the mental health problems and vulnerabilities of children and young people during the early phase of the COVID-19 pandemic using a secondary analysis of survey data collected from mental health care professionals in the UK (Johnson et al., 2020). Specifically, we examined professionals’ views on which groups of children and young people they are most concerned about and what they thought were the most challenging problems for this population during the early phase of the pandemic, as well as their views on new mental health problems arising as a result of the pandemic.

## 2. Methods

A secondary analysis of an online survey of mental health professionals in the UK conducted during an early phase of the pandemic (between 22 April 2020 and 12 May 2020). The full methods of the primary study are reported here (Johnson et al., 2020), including how the survey was developed and how participants were recruited. The King’s College London research ethics committee approved this study (MRA-19/20-18372). In summary, the survey comprised closed- and open-ended questions on: work-related challenges experinced during the pandemic, sources of support in managing work-related challenges, and persepctives on the needs of services users and family carers. The survey was disseminated through a range of channels, including professional networks, social media, and voluntary sector organsiations. Attempts were made to disseminate and make the survey inclusive of professionals from Black and minoritized ethnic groups. After being asked general questions on roles, work-related challenges encountered during COVID-19, sources of help in managing these problems, and their perception of problems faced by service users and family carers, survey branching was used to ask specific questions to staff from specific settings and specialities. These questions included changes to practice during COVID-19, the perceived benefits and limitations of these changes, and views on whether it would be beneficial to retain changes after COVID-19. Participants were included in the main analysis who reported working with children and adolescents, either exclusively or in combination with other service user groups. This resulted in a final sample of *N* = 601 professionals.

To address the research questions, the present analysis included 25 survey questions, 23 of which asked professionals the extent to which different problems were relevant among service users and carers they were in contact with (i.e., ‘Since mid-March 2020, how relevant do you think each of the following problems are among the service users and carer you are currently in contact with?’), with responses rated on a five-point scale from ‘Not relevant’ to ‘Extremely relevant’. These problems included loneliness (‘Loneliness due to or made worse by social distancing, self-isolation and/or shielding), access to mental health care (‘Lack of access to usual support from NHS mental health services’), and access to other services (‘Lack of access to usual support from other services (primary care, social care, voluntary sector)’). These responses were analysed using descriptive statistics.

Two free-text survey questions on vulnerabilities (‘Are there any groups of clients about whom you are particularly concerned at present? Please tell us about this:’) and mental health problems (‘Are you seeing any mental health problems that seem to arise directly from the current pandemic? If so, please tell us about them:’) were analysed using thematic analysis, involving familiarization (reading and re-reading the responses), coding (assigning descriptors to the data), generating themes (identifying patterns and grouping the codes), reviewing themes (ensuring the themes appropriately represent the data), defining and naming themes, and writing up (Braun & Clarke, 2006). A copy of the survey is available at this web address: https://opinio.ucl.ac.uk/s?s=67819.Demographic characteristics of study participants are shown in Table 1.

**Table 1.** Demographic characteristics of study participants

|  | *n* | % |
| --- | --- | --- |
| **Sector** |  |  |
| NHS | 491 | 82 |
| Social care | 36 | 6 |
| Voluntary | 51 | 8 |
| Community | 15 | 3 |
| Private | 35 | 6 |
| **Setting** |  |  |
| Inpatient | 119 | 20 |
| Crisis house | 7 | 1 |
| Residential | 12 | 2 |
| Crisis assessment | 112 | 19 |
| CMHT | 364 | 61 |
| Community | 48 | 8 |
| Other | 112 | 19 |
| **Profession** |  |  |
| Clinical or counselling psychologist | 105 | 17 |
| Nurse | 149 | 25 |
| Occupational therapist | 26 | 4 |
| Other qualified therapist | 68 | 11 |
| Peer support worker | 13 | 2 |
| Psychiatrist | 50 | 8 |
| Social worker | 37 | 6 |
| Manager or no professional qualification | 24 | 4 |
| Other work | 129 | 21 |
| Management responsibilities | 359 | 60 |
| Time worked (years) (M, SD) | 13.70 | 10.22 |
| **Region** |  |  |
| England | 496 | 83 |
| Northern Ireland | 10 | 2 |
| Scotland | 47 | 8 |
| Wales | 24 | 4 |
| Other | 8 | 1 |
| Not reported | 16 | 3 |

*N* = 601.

## 3. Results

Descriptive statistics of the problems experienced by service users reported by professionals are shown in Table 2, and key findings are discussed with the qualitative data analysis.

**Table 2.**
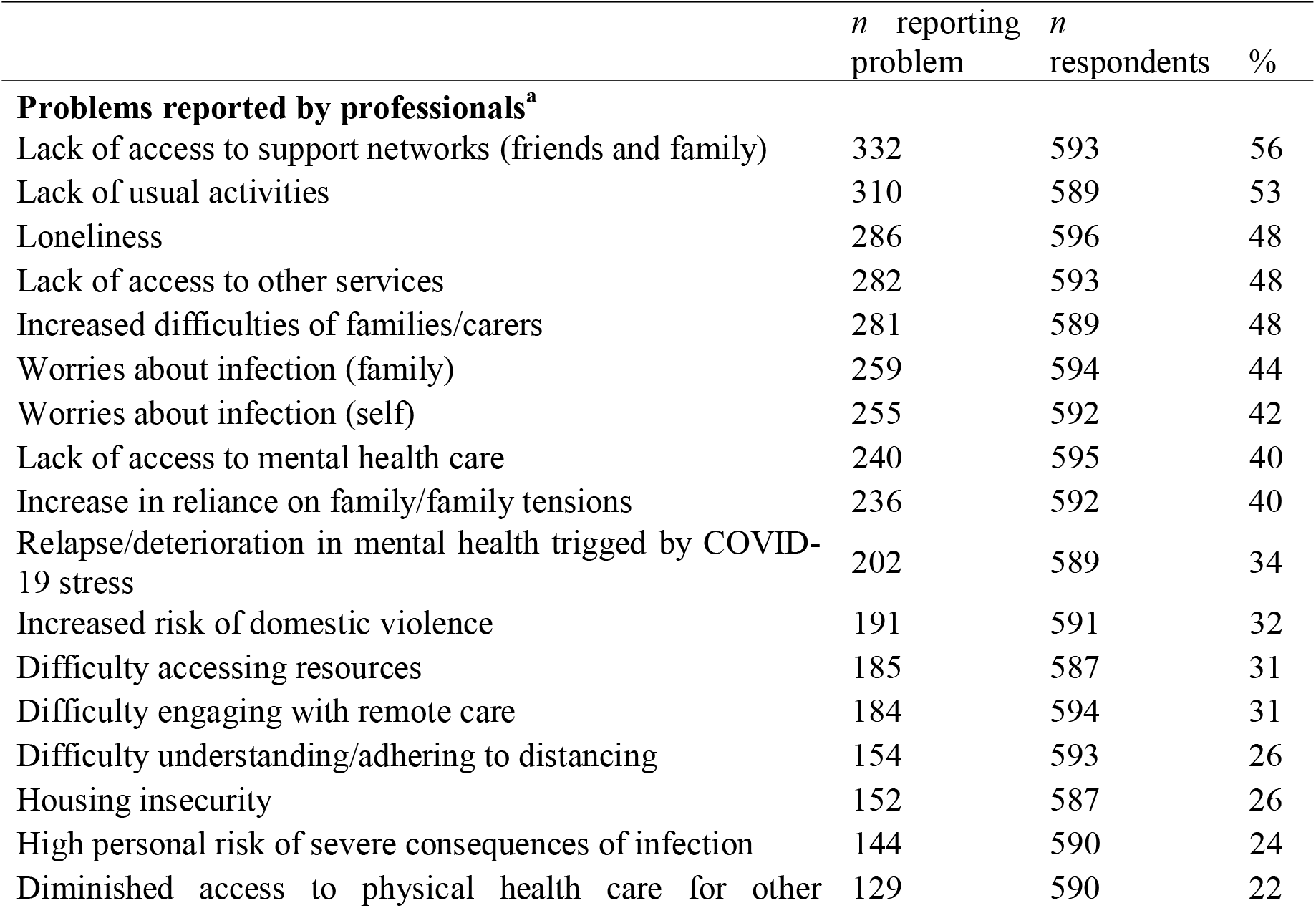
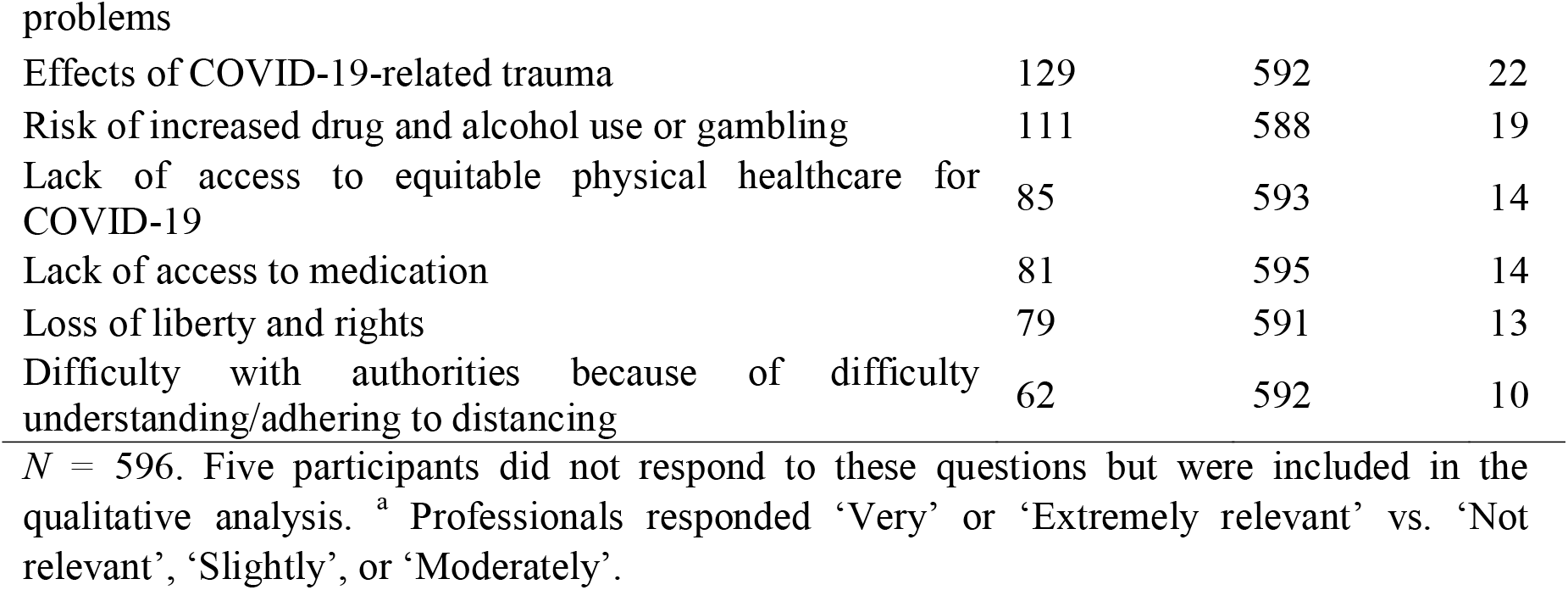
Problems experienced by service users reported by professionals.

Overall, four themes were found in the data in relation to groups about whom professionals were concerned and new problems arising from the pandemic (see Table 3). For the groups about whom professionals are concerned, three areas were found: young people experiencing pre-existing social inequalities, young people with limited access to support and services, and young people with complex or co-morbid mental and physical health issues. For new problems arising from the pandemic, one theme was found: an increase or exacerbation of mental health difficulties and challenging behaviours.

**Table 3.** Question and themes

| Question | Themes |
| --- | --- |
| Groups about whom professional are concerned | Exacerbation of pre-existing social inequalities |
|  | Young people with limited access to support and services |
|  | Young people with complex or co-morbid mental and physical health issues |
| New problems arising from the pandemic | An increase or exacerbation of mental health difficulties and challenging behaviours |

### 3.1 Groups about whom professionals are concerned

#### 3.1.1 Exacerbation of pre-existing social inequalities

Professionals were concerned about young people experiencing economic disadvantage and young people from minority ethnic groups, as pre-existing social inequalities result in increased risk of infection and serious infection due to increased likelihood of pre-existing physical health problems, crowded or unstable housing, and limited support. Overall, 31% of professionals reported concerns about difficulty accessing resources and 26% about housing insecurity. Limited access to food and free school meals were of particular concern, as were families with no stable income and the corresponding additional tensions of exacerbated financial difficulties meaning access to basic resources may become more challenging.

> *“Children at risk due to parent/carer job losses and lack of basic provisions”* (all quotes are from professionals’ responses)

Families experiencing economic disadvantage were also mentioned as experiencing additional barriers to accessing outdoor spaces and remote education. Professionals raised concerns about young people living in confined, crowded, or chaotic living arrangements, including those in hospital where leave, visits, and community-transitions may be reduced or unavailable and others requiring admission where availability may be reduced, for example:

> *“young people suffering from acute mental health conditions who are inpatients in medium-secure forensic settings. Some of them had been moving forward to transition back to the community and on a graduated plan of leave back to their family/community and discharge. This has now been put on hold because of lockdown and risk of introducing virus into the unit, and therefore risk to staff and service users alike.”*

Similarly, young people in hospital, care, or secure settings may have their daily routines drastically changed and periods of leave or discharge put on hold. Homeless young people and those in temporary or unstable housing were described as particularly challenging to reach, with reduced access to food and support due to closures.

Families with social care needs or experiencing domestic violence or abuse were another group for whom professionals were concerned, due to additional time at home when stressors and family tensions may be exacerbated and there are fewer monitoring and support opportunities e.g., by schools and services. Indeed, 32% of professionals reported concerns related to increased risk of domestic violence.

> *“Young people in unsafe homes where they cannot be seen or heard talking to external organisations or seeking mental health support”*

Children with pre-existing social care needs (either above or below threshold for receiving social care services) now not attending school, or those awaiting assessment or care, were particularly concerning to professionals.

Professionals described concerns about families with caring needs, primarily those with caring responsibilities with reduced access to respite care. This was compounded by limited available activities and additional stressors for the carer and those being cared for. Correspondingly, professionals were concerned about managing challenging behaviour being more difficult

> *“Carer stress and burden because of a lack of respite - i.e. support workers and activities for the young person”*

### 3.1.2 Young people with limited access to support and services

This group included young people lacking supportive family or with challenging family relationships, which may be exacerbated by the increased tensions and amount of time together and the lack of access to safe spaces, key sources of support, and activities that help to regulate emotions and behaviour, such as seeing friends or wider family members, school, outdoor activities, exercise, and access to therapy. The increased reliance on family and family tensions were reported as a concern by 40% of professionals. Young people living alone were described as a source of concern due to isolation and limited access to practical support from family and friends. Almost half (48%) of professionals reported concerns about loneliness and over half (56%) reported concerns about lack of access to support networks. Similarly, young people with family overseas, or with only limited access to or use of social media, were described as facing increased isolation. A lack of social support was described by professionals as posing an important mental health risk to young people generally and young people who had previously been experiencing improvement or remission of mental health difficulties (prior to the start of the pandemic) in particular.

Lack of usual activities was reported by 53% of professionals as a concern. Not attending school and reduced social activities were described as a double-edged sword for young people for whom these activities are challenging, such as those with school attendance and social anxiety problems, in that the improvement of mental health due to reduction of these stressors may be off-set by increased difficulties with reintegration when restrictions are lifted. Similarly, there were reports of young people with pre-existing mental health difficulties experiencing improvement because of reduced social stressors, no longer feeling disadvantaged in function relative to others, and building up confidence leaving the house as places were quieter (in the case of social anxiety). Nevertheless, there were concerns about expected increased levels of depression, anxiety, OCD, and agoraphobia in the long-term.

Professionals raised concerns about young people and families not accessing support and/or medications due to restrictions, shielding, concerns over catching or transmitting the virus, a lack of access to remote care, or difficulties engaging with remote care. The loss of support networks through accessing services and support groups, and the role these play in preventing relapse, were highlighted. In the quantitative responses, 40% of professionals reported concerns about lack of access to mental health services, and 7% reported concerns about lack of access to other services. The role of schools was mentioned, regarding young people who had been accessing mental health support in schools but were suddenly not able to.

> *“Children and young people who accessed counselling in schools who can’t access services from home”*

The lack of physical and mental healthcare for those with eating disorder was highlighted, and more generally 22% of professionals reported concerns related to diminished access to physical health care for other problems.

> *“Eating disorder patients or those without an ED but who are extremely low weight without access to Paediatric provision”*

Providing care remotely was described as positive but insufficient in reaching all those in need and especially traditionally underserved groups. Moreover, it was difficult to provide the same level of care remotely as was provided face-to-face, as was ensuring access for certain groups, including young people for whom English is not a first language, deaf young people, and young people who experience different realities.

> *“Really worried about psychotic young people. Might make them more paranoid than usual and trigger a deterioration in their mental state.”*
>
> *“People who experience different realities around government surveillance, the screen talking to them etc who can’t use tech/video calls”*

Providing substance use services without face-to-face contact was particularly mentioned as a concern. Finally, concern was raised about young people with self-harm, especially as pandemic guidance in the early stages advised against attending Accident and Emergency care.

### 3.1.3 Young people with complex or co-morbid mental and physical health issues

Overall, 34% of professionals reported concerns about relapse or deterioration in mental health triggered by COVID-19 stress. Dual concerns were raised for young people whose mental health difficulties had been improving and might now experience deterioration or relapse due to additional stressors.

> *“young people with autism and relapse in mental health conditions - sometimes resulting in increased family tension and aggression from the young person”*

The potential role of long-term medication use on physical health was described as a concern in terms of increase risk of infection or severe infection. Professionals described concerns for young people with particular pre-existing mental health needs, including generalized anxiety, health anxiety, OCD, self-harm or suicidality, trauma, and psychosis. Concerns were raised about young people with eating disorder or substance use problems and increased risk of infection or severe infection due to impaired physical health.

Young people with neurodevelopmental difficulties such as autism spectrum conditions, ADHD, and learning disabilities, were also highlighted as a vulnerable group by professionals. Primarily, concerns pertained to the sudden loss of structure afforded by attending school, disrupted access to education, and, more generally, changes to the environment and the lack of the usual daily routine.

> *“Children and young people with a mental health disorder and co-morbid learning disability, often with associated challenging behaviours. No usual school routine or programmed activities and families with limited resources”*
>
> *“I am concerned about children with neurodevelopmental difficulties whose parents and families are under greater stress due to the lack of the usual respite facilities. Respite care is generally inadequate anyway, but currently the situation is far worse”*

Limited access to outdoor spaces, recreational activities, and exercise were described as particularly concerning given the important role such activities play in providing sensory needs input and regulating mood. Corresponding deterioration in mood was described, resulting in additional behavioural difficulties and strain on the family. Remote services were described as being less accessible to young people with neurodevelopmental difficulties, and barriers to understanding the pandemic and health and safety practices were described as additional stressors. Difficultly engaging with remote care was reported as a concern by 31% of professionals in the quantitative questions.

### 3.2 New problems arising from the pandemic

New mental health problems arising from the pandemic were reported by professionals as arising from stress and worry about the self and others directly related to the virus; drastically altered routines and structures; concerns about pre-existing health conditions; financial concerns; loneliness, social isolation, and withdrawal; frustration with physical distancing; carer stress due to not being able to access and care for loved ones; a lack of respite care; increased family tensions due to increased time with each other at home; and lack of access to culturally significant products or activities. Although the exacerbation of pre-existing mental health problems and new difficulties for those with other pre-existing mental health problems were described, onset of mental health problems in those with no previous history was discussed equally.

> *“Triggers for mental health relapse in people currently known to services*.
>
> *Depression/anxiety/suicidal ideation and actual suicide attempts by people not previously known to services”*

#### 3.2.1 Increase or exacerbation of mental health difficulties and challenging behaviours

Stress, worry, generalized anxiety disorder, health anxiety, and OCD with a focus on contamination anxiety, were commonly reported. Increased levels of depression and anxiety were described with associated lack of motivation, fear of the future, hopelessness, poor sleep, irritability, and lack of self-care behaviours. For example,

> *“High rates in anxiety (not just health anxiety) but anxieties regarding sense of control, understanding the future, fear of what this means”*
>
> *“Parental ability to nurture relationships with young people due to their own mental health being impacted”*.

Specific anxieties were mentioned, such as parent/carer anxiety, birthing anxiety, access to food, and fear of the police and State control (in light of increased restrictions), but the most commonly reported anxieties by professionals were young people’s concerns about their future and post-restriction reintegration.

There were general reports of increased levels of aggression and challenging behaviour pertaining to increased anxiety or emotion dysregulation, which at times was mentioned particularly for young people with neurodevelopmental difficulties. There were some reports of increased substance use and a small number of reports of increased gambling and gaming addiction. Professionals reported more Mental Health Act assessments being conducted and more individuals being detained under mental health legislation, including those with no previous history of mental health difficulties; in some instances, detention was precipitated by behavioural difficulties or increased levels of substance use and risky behaviours leading to police involvement.

> *“Yes. More S136 [Section 136 used when there are serious concerns mental health needs or safety] and since the extension to the lockdown more acuity within in the inpatient units. More reckless behaviours such as drug taking and alcohol misuse resulting in S136”*

Increased family stress, conflict, and domestic violence and abuse were reported. Increases in self-harm and suicidality were mentioned in relation to new onset, relapse despite long periods of remissions, and young people with pre-existing mental health needs.

Trauma was described less often but referred to increased experiences of hypervigilance, bereavement, unresolved grief due to a lack of mourning, and reactivation of existing trauma symptoms. On a more general level, professionals also reported prolonged birth trauma due to a lack of partner and family attendance. Lastly, there were reports of exacerbation of eating disorders or relapse but not necessarily new onset:

> *“Also, for patients with anorexia nervosa they are struggling to cope with shopping and food availability which is already highly problematic for them. Many are reducing their food intake due to not being able to access their [safe foods]”)*.

## 4. Discussion

Through the secondary data analysis of survey data investigating mental health professionals’ perspectives of the pandemic on mental health services, the present study examines their concerns about groups of children and young people more likely to be exposed to difficulties during the early phase of pandemic (Johnson et al., 2020). The study also addresses new mental health problems and recent challenging behaviours experienced by children and young people due to the pandemic.

Overall, professionals expressed concerns about limited access to support and services for children and young people. Such difficulties were noted in other studies as very relevant across all service user groups (Perrin et al., 2020; Radez et al., 2020; Singh et al., 2020). Alongside reduced service access, the deterioration of pre-existing mental health problems in the pandemic period were also addressed as a possible negative impact for various service users. This is consistent with other studies that have found children with anxiety, ADHD, and special educational needs have reported an increase in symptoms and have reduced access to support as a result of capacity and service restructuring (Barnardo’s, 2020). The increase in symptoms are likely exacerbated by additional stressors, including increased hardships within the families (e.g., job loss or income reduction) or additional caregiving responsibilities (Gassman-Pines et al., 2020; Goldman et al., 2020; Silva Junior et al., 2020).

Our findings specifically captured that the fewer monitoring and support opportunities through schools and services during the lockdown brought increased concerns for young people with pre-existing mental health needs. Relatedly, reduced access to already strained and under-funded children and young people’s mental health services (CYPMHS) during the pandemic as a result of high demand and service restructuring has left many without access to treatment. Providing relevant interventions for children and young people in early stages is vital to prevent problems from becoming more serious and entrenched (Goodman et al, 2002; Jones, 2013). Similarly, there has been a decrease in referrals to frontline staff (e.g., Barnardo’s) which is partially attributable to the lack of contact with teachers, support staff, or other community groups who typically report cases of abuse and neglect. Also, the results suggest children and young people have been particularly impacted by pre-existing social inequalities during the pandemic. Professionals reflected on the increased likelihood of being exposed to various difficulties for children and young people from low socio-economic status and minority ethnic background, including exacerbated financial difficulties at home and limited access to free school meals due to the school closures. These findings complement other studies that have raised particular concerns about young people living in disadvantaged living arrangements and the subsequent impact on their mental health (Endale et al., 2020; Galvin & Kaltner, 2020; Pierce et al., 2020). Echoing previous studies, professionals reported some children and young people have found relief from social stressors during the pandemic. However, for those with social anxiety, government mandates to isolate may become positive reinforcements, and reintegration may instigate a spike in anxiety once restrictions ease (Morrissette, 2021). Despite the short-term lessening of symptoms, it is proposed that clinicians continue to treat social anxiety disorder and incorporate virtual exposure to social situations.

### 4.1 Strengths and limitations

This study helps shed light on a novel situation that is still unfolding and offers insight into the challenges many children and young people are facing. One strength of this study is the diversity of professionals who completed the survey. This may help paint a fuller picture on which groups of children and young people may be suffering disproportionally and identify those at risk of developing symptoms stemming from stress and worry. Additionally, this data was collected during the height of the first lockdown, allowing us to gain insight into professionals’ concerns during the most restrictive phase of the pandemic. This may be particularly relevant as communities face subsequent lockdowns or during future pandemics so contingency plans can be put in place to mitigate against the negative mental health consequences.

Several limitations should also be considered. First is the use of the convenience sample to collect survey data which was used in this analysis. As such, findings may not be representative of the professional population at large as there may be some underrepresented groups such as community organisations, health visitors, children’s nurses, and teachers who are likely to know young people and families intimately and to capture stronger opinions than what is representative of the entire workforce. Indeed, as the pandemic emerged, many job roles shifted to cope with the increased demand on CYPMHS and social services. Next, the sample included professionals who see a range of age groups and we therefore cannot be certain their responses are in reference to children and young people. To mitigate this, we aimed to include quotes that explicitly mentioned children and young people. Lastly, the study sample had a poor representation of staff from minority ethnic groups despite the original study’s efforts during recruitment to improve this (Johnson et al., 2020).

### 4.2 Implications and Future Research

The increasing professional concern about particularly vulnerable groups of children and young people and the growing number of mental health issues brings about clinical and public health implications. As restrictions ease and children and young people return to school, some may have particular anxieties relating to feeling safe or mixing with peers. Upskilling teachers, parents, and those working with young people to spot the signs of difficulties and signpost to appropriate support is crucial for early intervention. However, the reduced access to services means there are a large number of young people in need of self-management strategies and/or professional support. Online resources and interventions can play a great role in increasing access, as well as practitioners engaging in tele-psychiatry or tele-neuropsychology (Hasking et al., 2021; Patra & Patro, 2020; Peterson et al., 2021). Schools may also consider including more mental health lessons and self-care activities in their curriculum.

Throughout the pandemic there has been an overwhelming response from authors, academics, and teachers to produce materials to help support young people at this incredibly challenging time. Research remains crucial going forward in order to have a clear picture of the ongoing mental health needs for children and young people so support and funding can be optimised. Future research must include the views of children, young people, and their families to better understand the impact of the pandemic (Branquinho et al., 2020). Researchers may want to focus on young people from minority ethnic groups, those in or leaving care settings, and the LGBTQ+ community, as there is evidence they are disproportionately affected. With schools being a place where mental health provision is frequently employed, the views of educational professionals may also be insightful. Given the descriptive nature of this study and the previous study from which this data was drawn, future research may also wish to explore associations between different predictor variables (e.g., sector, setting, or professional grouping) which could help direct resources and support if differences are found.

Lived experience commentary: Ellie Brooks-Hall

The conclusions drawn from the research resonated with my personal experience of the pandemic and my understanding of my peers’ experiences. Particularly for young people, having limited access to their usual ‘distractors’ or coping mechanisms has been a major struggle over the last year, as well as the loneliness caused by not being at school or able to socialise to provide some escapism, so I was very happy to see these themes raised amongst the findings.

I also was grateful to see their suggestion of ‘mental health lessons and self-care activities’ in schools, as caring for mental health is a widely dismissed area of education. Mental health challenges are still seen as an ‘excuse’ but including mental health care in the curriculum is an important first step to becoming an accepting and educated school population. However, there are concerns with this suggestion such as a potential promotion of a medical model that disregards social determinants of health.

A number of the clinicians involved in this study were seeing adults as well as children and young people, and I question whether all their responses were about children and young people or whether they were extrapolating experiences from other demographics. Furthermore, the data in the study was collected quickly during the first lockdown in England, limiting the conclusions we can draw about the experiences of young people at different times and places. Young people experienced more issues when starting to come out of lockdown, from the anxiety and uncertainty about post-lockdown life, socialising again, and whether this will cause an increase in cases of COVID-19. We have also subsequently had repeated lockdowns with more of an impact on mental health than in the first lockdown when this data was collected.

While this study is useful to understand the problems of the first lockdown, further research is needed to explore the continued impact for children and young people, and their families, including in comparison to their experiences before the pandemic. Young people with various lived experiences must be involved throughout the entire process for future research to ensure both the questions and findings are relevant to their experience.

## Data Availability

The survey dataset is currently being used for additional research by the author research group and is therefore not currently available in a data repository. A copy of the survey is available at this web address: https://opinio.ucl.ac.uk/s?s=67819.

## Acknowledgements

List of The COVID-19 Mental Health Policy Research Unit Group members: Andy Bell (Centre for Mental Health, London, UK), Francesca Bentivegna (NIHR Mental Health Policy Research Unit, Division of Psychiatry, University College London, London, UK), Joseph Botham (NIHR Mental Health Policy Research Unit, Institute of Psychiatry, Psychology & Neuroscience, King’s College London, London, UK), Sarah Carr (School of Social Policy/Institute for Mental Health, University of Birmingham, Birmingham, UK), Una Foye (NIHR Mental Health Policy Research Unit), Steve Gillard (Population Health Research Institute, St George’s University of London, London, UK), Lucy Goldsmith (Population Health Research Institute, St George’s, University of London, London, UK), Lisa Grünwald (NIHR Mental Health Policy Research Unit, Division of Psychiatry, University College London, London, UK; North East London NHS Foundation Trust, London, UK), Jasmine Harju-Seppänen (NIHR Mental Health Policy Research Unit, Division of Psychiatry, University College London, London, UK; Division of Psychology and Language Sciences, University College London, London, UK), Stephani Hatch (NIHR Mental Health Policy Research Unit, Institute of Psychiatry, Psychology & Neuroscience, King’s College London, London, UK), Claire Henderson (NIHR Mental Health Policy Research Unit, Institute of Psychiatry, Psychology & Neuroscience, King’s College London, London, UK), Louise Howard (NIHR Mental Health Policy Research Unit, Institute of Psychiatry, Psychology & Neuroscience, King’s College London, London, UK), Tamar Jeynes (Division of Psychiatry (NIHR Mental Health Policy Research Unit COVID-19 Co-Production Group), University College London, London, UK), Helen Killaspy (Division of Psychiatry, NIHR Mental Health Policy Research Unit, University College London, London, UK; Camden and Islington NHS Foundation Trust, London, UK), Sabine Landau (NIHR Mental Health Policy Research Unit), Rebecca Lane (Anna Freud Centre, London, UK), Sarah Ledden (NIHR Mental Health Policy Research Unit, Division of Psychiatry, University College London, London, UK), Monica Leverton (NIHR Mental Health Policy Research Unit, Division of Psychiatry, University College London, London, UK), Brynmor Lloyd-Evans (Division of Psychiatry, NIHR Mental Health Policy Research Unit, University College London, London, UK), Jo Lomani (Population Health Research Institute, St George’s, University of London, London, UK), Natasha Lyons (NIHR Mental Health Policy Research Unit, Division of Psychiatry, University College London, London, UK), Paul McCrone (Faculty of Education, Health and Human Sciences, University of Greenwich, London, UK), Chukwuma U Ntephe (NIHR Mental Health Policy Research Unit, Division of Psychiatry, University College London, London, UK; South London and Maudsley NHS Foundation Trust, London, UK), Josephine Enyonam Ocloo (NIHR Mental Health Policy Research Unit, Institute of Psychiatry, Psychology & Neuroscience, King’s College London, London, UK), Sian Oram (NIHR Mental Health Policy Research Unit), David Osborn (NIHR Mental Health Policy Research Unit, Division of Psychiatry, University College London, London, UK; Camden and Islington NHS Foundation Trust, London, UK), Alexandra Papamichail (NIHR Mental Health Policy Research Unit), Steve Pilling (Division of Psychology and Language Sciences, University College London, London, UK), Konstantina Poursanidou (Division of Psychiatry (NIHR Mental Health Policy Research Unit COVID-19 Co-Production Group), University College London, London, UK), Rachel Rowan Olive (Division of Psychiatry (NIHR Mental Health Policy Research Unit COVID-19 Co-Production Group), University College London, London, UK), Hannah Rachel Scott (NIHR Mental Health Policy Research Unit, Division of Psychiatry, University College London, London, UK), Prisha Shah (Division of Psychiatry (NIHR Mental Health Policy Research Unit COVID-19 Co-Production Group), University College London, London, UK), Luke Sheridan Rains (Division of Psychiatry, NIHR Mental Health Policy Research Unit, University College London, London, UK), Thomas Steare (NIHR Mental Health Policy Research Unit, Division of Psychiatry, University College London, London, UK), Ruth Stuart (NIHR Mental Health Policy Research Unit, Institute of Psychiatry, Psychology & Neuroscience, King’s College London, London, UK), André Tomlin (National Elf Service Ltd, Oxford, UK), Kati Turner (Population Health Research Institute, St George’s, University of London, London, UK), and Vasiliki Tzouvara (Department of Mental Health Nursing, Florence Nightingale Faculty of Nursing, Midwifery & Palliative Care, King’s College London, London, UK).

## References

Andrew, A., Cattan, S., Costa-Dias, M., Farquharson, C., Kraftman, L., Krutikova, S., Phimister, A., & Sevilla, A. (2020). Learning during the lockdown: real-time data on children’s experiences during home learning. Ifs.

Barnardo’s. Time for a Clean Slate; Children’s Mental Health at the Heart of Education. https://www.barnardos.org.uk/sites/default/files/uploads/time-for-clean-slate-mental-health-at-heart-education-report.pdf (2020).

Benson-Allott, C. Out of sight? Vulnerable Young People: COVID-19 Response. COVID 19 Response Final Report (2020) doi:10.1525/FQ.2011.65.2.14.

Blake-Holmes, K. (2020). Understanding the needs of young carers in the context of the COVID-19 global pandemic. Norwich: University of East Anglia.

Branquinho, C., Kelly, C., Arevalo, L. C., Santos, A., & Gaspar de Matos, M. (2020). “Hey, we also have something to say”: A qualitative study of Portuguese adolescents’ and young people’s experiences under COVID-19. Journal of Community Psychology, 48(8), 2740–2752. https://doi.org/10.1002/jcop.22453

Braun, V., & Clarke, V. (2006). Using thematic analysis in psychology. Qualitative Research in Psychology, 3(2), 77–101. https://doi.org/10.1191/1478088706qp063oa

Children’s Commissioner for Wales. Coronavirus and Me. https://www.childcomwales. org.uk/wp-content/uploads/2020/06/FINAL_ formattedCVRep_EN.pdf (2020).

Cortina, M. A., Gilleard, A., & Deighton, J. (2020). Emerging evidence: coronavirus and children and young people’s mental health. Evidence Based Practice Unit, London.

Cortina, M. A., Gilleard, A., Deighton, J. & Edbrooke-Childs, J. (2020). Emerging evidence (Issue 2): coronavirus and children and young people’s mental health. Evidence Based Practice Unit, London.

Das, N. (2020). Psychiatrist in post-COVID-19 era – Are we prepared? Asian Journal of Psychiatry, 51, 102082. https://doi.org/10.1016/j.ajp.2020.102082

Endale, T., St Jean, N., & Birman, D. (2020). COVID-19 and refugee and immigrant youth: A community-based mental health perspective. Psychological Trauma: Theory, Research, Practice, and Policy. https://doi.org/10.1037/tra0000875

Galvin, M., & Kaltner, M. (2020). Understanding the Impact of COVID-19 on Out-of-Home Care in Australia. EY. https://assets.ey.com/content/dam/ey-sites/ey-com/en_au/topics/covid-19-response/ey-impacts-of-covid-19-on-oohc-.pdf

Gassman-Pines, A., Ananat, E. O., & Fitz-Henley, J. (2020). COVID-19 Crisis Impacts on Parent and Child Psychological Well-being. Pediatrics, 146(4), e2020007294. https://doi.org/10.1542/peds.2020-007294

Gilleard, A., Lereya, S. T., Tait, N., Edbrooke-Childs, J., Deighton, J., & Cortina, M. A. (2020). Emerging evidence (Issue 3): coronavirus and children and young people’s mental health. Evidence Based Practice Unit, London.

Goldman, P. S., van Ijzendoorn, M. H., Sonuga-Barke, E., & Lancet Institutional Care Reform Commission Group (2020). The implications of COVID-19 for the care of children living in residential institutions. The Lancet. Child & adolescent health, 4(6), e12. https://doi.org/10.1016/S2352-4642(20)30130-9

Goodman, R., Ford, T., & Meltzer, H. (2002). Mental health problems of children in the community: 18 month follow up. BMJ (Clinical research ed.), 324(7352), 1496–1497. https://doi.org/10.1136/bmj.324.7352.1496

Graell, M., Morón-Nozaleda, M. G., Camarneiro, R., Villaseñor, Á., Yáñez, S., Muñoz, R., Martínez-Núñez, B., Miguélez-Fernández, C., Muñoz, M., & Faya, M. (2020). Children and adolescents with eating disorders during COVID-19 confinement: Difficulties and future challenges. European Eating Disorders Review, 28(6), 864–870. https://doi.org/10.1002/erv.2763

Hasking, P., Lewis, S. P., Bloom, E., Brausch, A., Kaess, M., & Robinson, K. (2021). Impact of the COVID-19 pandemic on students at elevated risk of self-injury: The importance of virtual and online resources. School Psychology International, 42(1), 57–78. https://doi.org/10.1177/0143034320974414

Hawke, L. D., Monga, S., Korczak, D., Hayes, E., Relihan, J., Darnay, K., Cleverley, K., Lunsky, Y., Szatmari, P., & Henderson, J. (2020). Impacts of the COVID-19 pandemic on youth mental health among youth with physical health challenges. Early Intervention in Psychiatry, 2020, 1–8. https://doi.org/10.1111/eip.13052

Institute of Health Equity. Health Equity in England: The Marmot Review 10 Years On. Available online at: http://www.instituteofhealthequity.org/about-us/the-institute-of-health-equity/our-current-work/collaborating-with-the-health-foundation- (Accessed February 2020).

Johnson, S., Dalton-Locke, C., Vera San Juan, N., Foye, U., Oram, S., Papamichail, A., Landau, S., Rowan Olive, R., Jeynes, T., Shah, P., Sheridan Rains, L., Lloyd-Evans, B., Carr, S., Killaspy, H., Gillard, S., & Simpson, A. (2020). Impact on mental health care and on mental health service users of the COVID-19 pandemic: a mixed methods survey of UK mental health care staff. Social Psychiatry and Psychiatric Epidemiology. https://doi.org/10.1007/s00127-020-01927-4

Jones, P.B. (2013). Adult mental health disorders and their age at onset. The British Journal of Psychiatry, 202(54), s5–s10

Liu, C.H., Stevens, C., Conrad, R. C., & Hahm, H. C. (2020). Evidence for elevated psychiatric distress, poor sleep, and quality of life concerns during the COVID-19 pandemic among U.S. young adults with suspected and reported psychiatric diagnoses. Psychiatry Research, 292(January). https://doi.org/10.1016/j.psychres.2020.113345

Liu, Cindy H., & Doan, S. N. (2020). Psychosocial Stress Contagion in Children and Families During the COVID-19 Pandemic. Clinical Pediatrics, 59(9–10), 853–855. https://doi.org/10.1177/0009922820927044

Liu, Cindy H., Zhang, E., Wong, G. T. F., Hyun, S., & Hahm, H. “Chris.” (2020). Factors associated with depression, anxiety, and PTSD symptomatology during the COVID-19 pandemic: Clinical implications for U.S. young adult mental health. Psychiatry Research, 290(June). https://doi.org/10.1016/j.psychres.2020.113172

Morrissette, M. (2021). School Closures and Social Anxiety During the COVID-19 Pandemic. Journal of the American Academy of Child & Adolescent Psychiatry, 60(1), 6–7. https://doi.org/10.1016/j.jaac.2020.08.436

Nonweiler, J., Rattray, F., Baulcomb, J., Happé, F., & Absoud, M. (2020). Prevalence and Associated Factors of Emotional and Behavioural Difficulties during COVID-19 Pandemic in Children with Neurodevelopmental Disorders. Children, 7(9). https://doi.org/10.3390/children7090128

Patra, S., & Patro, B. K. (2020). COVID-19 and the need for child and adolescent telepsychiatry services, a case report. Asian Journal of Psychiatry, 54, 102298. https://doi.org/10.1016/j.ajp.2020.102298

Perrin, P. B. et al. Rapid telepsychology deployment during the COVID-19 pandemic: A special issue commentary and lessons from primary care psychology training. J. Clin. Psychol. 76, 1173–1185 (2020).

Peterson, R. K., Ludwig, N. N., & Jashar, D. T. (2021). A case series illustrating the implementation of a novel tele-neuropsychology service model during COVID-19 for children with complex medical and neurodevelopmental conditions: A companion to Pritchard et al., 2020. Clinical Neuropsychologist, 35(1), 99–114. https://doi.org/10.1080/13854046.2020.1799075

Pierce, M., Hope, H., Ford, T., Hatch, S., Hotopf, M., John, A., Kontopantelis, E., Webb, R., Wessely, S., McManus, S., & Abel, K. M. (2020). Mental health before and during the COVID-19 pandemic: a longitudinal probability sample survey of the UK population. The Lancet Psychiatry, 0366(20), 1–10. https://doi.org/10.1016/S2215-0366(20)30308-4

Radez, J., Reardon, T., Creswell, C., Lawrence, P. J., Evdoka-Burton, G., & Waite, P. (2021). Why do children and adolescents (not) seek and access professional help for their mental health problems? A systematic review of quantitative and qualitative studies. European child & adolescent psychiatry, 30(2), 183–211. https://doi.org/10.1007/s00787-019-01469-4

Silva Junior, F. J. G. Da, Sales, J. C. E. S., Monteiro, C. F. D. S., Costa, A. P. C., Campos, L. R. B., Miranda, P. I. G., … Lopes-Junior, L. C. (2020). Impact of COVID-19 pandemic on mental health of young people and adults: A systematic review protocol of observational studies. BMJ Open. http://doi.org/10.1136/bmjopen-2020-039426

Singh, S., Roy, D., Sinha, K., Parveen, S., Sharma, G., & Joshi, G. (2020). Impact of COVID-19 and lockdown on mental health of children and adolescents: A narrative review with recommendations. Psychiatry Research, 293, 113429. doi:10.1016/j.psychres.2020.113429

Tanir, Y.,Karayagmurlu, A., Kaya, İ., Kaynar, T. B., Türkmen, G., Dambasan, B. N., Meral, Y., & Coşkun, M. (2020). Exacerbation of obsessive compulsive disorder symptoms in children and adolescents during COVID-19 pandemic. Psychiatry Research, 293(July), 3–7. https://doi.org/10.1016/j.psychres.2020.113363

